## Supplementary file for "Eating Disorders In weight-related Therapy (EDIT): Protocol for a systematic review with individual participant data meta-analysis of eating disorder risk in behavioural weight management"

**Table S1: Search strategies**

| |
|---|
| **MEDLINE** |
| 1. exp Obesity/ |
| 2. exp Overweight/ |
| 3. obes*.tw. |
| 4. overweight.tw. |
| 5. 1 or 2 or 3 or 4 |
| 6. weight loss/ |
| 7. exp diet therapy/ |
| 8. exp bariatrics/ |
| 9. exp exercise/ |
| 10. anti-obesity agents/ or appetite depressants/ |
| 11. (diet* adj2 therap*).tw. |
| 12. bariatric*.tw. |
| 13. (low adj3 (energy or calor*) adj4 diet).tw. |
| 14. ((pharma* or diet* or obes* or lifestyle or behavio*) adj3 (interven* or treat* or therap*)).tw. |
| 15. ((calori* or diet*) adj3 (reduc* or restrict*)).tw. |
| 16. (weight adj3 (manag* or los*)).tw. |
| 17. (exercis* or physical activit*).tw. |
| 18. HAES.mp. |
| 19. health at every size.mp. |
| 20. (weight adj2 neutral).mp. |
| 21. nondiet.mp. |
| 22. (non adj2 diet).mp. |
| 23. (intuitive adj2 eat*).mp. |
| 24. mindful*.tw. |
| 25. 6 or 7 or 8 or 9 or 10 or 11 or 12 or 13 or 14 or 15 or 16 or 17 or 18 or 19 or 20 or 21 or 22 or 23 or 24 |
| 26. Body Image/ |
| 27. (body adj3 (accept* or dissatisf* or image or satisf* or appreciat* or esteem)).tw. |
| 28. "feeding and eating disorders"/ or anorexia nervosa/ or binge-eating disorder/ or bulimia nervosa/ or "feeding and eating disorders of childhood"/ |
| 29. (bulimi* adj3 symptom*).tw. |
| 30. (disorder* adj3 eat*).tw. |
| 31. (emotion* adj3 eat*).tw. |
| 32. (diet* adj3 restr*).tw. |
| 33. (binge adj3 eat*).tw. |
| 34. extreme weight loss.tw. |
| 35. loss of control.tw. |
| 36. drive for thinness.tw. |
| 37. ((weight or shape or eat*) adj3 concern).tw. |
| 38. (eat* adj2 behavi*).tw. |
| 39. 26 or 27 or 28 or 29 or 30 or 31 or 32 or 33 or 34 or 35 or 36 or 37 or 38 |
| 40. randomized controlled trial/ |
| 41. (randomi?ed controlled trial* or RCT* or (controlled adj3 trial)).mp. |
| 42. randomi?ed.ti. |
| 43. clinical trials as topic.sh. |
| 44. randomly.ab. |
| 45. trial.mp. |
| 46. clinical trial.mp. |
| 47. 40 or 41 or 42 or 43 or 44 or 45 or 46 |
| 48. 5 and 25 and 39 and 47 |
| **EMBASE** |

1. obesity/
2. obes*.tw.
3. overweight.tw.
4. 1 or 2 or 3
5. weight reduction/
6. diet therapy/ or diet restriction/ or low calory diet/ or low fat diet/
7. bariatric surgery/ or gastric banding/ or sleeve gastrectomy/
8. exercise/
9. antiobesity agent/
10. (diet* adj2 therap*).tw.
11. bariatric*.tw.
12. (low adj4 (energy or calor*) adj4 diet).tw.
13. ((pharma* or diet* or obes* or lifestyle or behavio*) adj3 (interven* or treat* or therap*)).tw.
14. ((calori* or diet*) adj3 (reduc* or restrict*)).tw.
15. (weight adj3 (manag* or los*)).tw.
16. (exercis* or physical activit*).tw.
17. HAES.mp.
18. health at every size.mp.
19. (weight adj2 neutral).mp.
20. nondiet.mp.
21. (non adj2 diet).mp.
22. (intuitive adj2 eat*).mp.
23. mindful*.tw.
24. 5 or 6 or 7 or 8 or 9 or 10 or 11 or 12 or 13 or 14 or 15 or 16 or 17 or 18 or 19 or 20 or 21 or 22 or 23
25. body image/
26. (body adj3 (accept* or dissatisf* or image or satisf* or appreciat* or esteem)).tw.
27. eating disorder/ or anorexia nervosa/ or binge eating disorder/ or bulimia/
28. feeding behavior/
29. (bulimi* adj3 symptom*).tw.
30. (disorder* adj3 eat*).tw.
31. (emotion* adj3 eat*).tw.
32. (diet* adj4 restrain*).tw.
33. (binge adj3 eat*).tw.
34. extreme weight loss.tw.
35. loss of control.tw.
36. drive for thinness.tw.
37. ((weight or shape or eat*) adj3 concern).tw.
38. 25 or 26 or 27 or 28 or 29 or 30 or 31 or 32 or 33 or 34 or 35 or 36 or 37
39. randomized controlled trial/ or controlled clinical trial/
40. (randomi?ed controlled trial* or RCT* or (controlled adj3 trial)).mp.
41. randomi?ed.ti.
42. randomly.ab.
43. trial.mp.
44. clinical trial.mp.
45. 39 or 40 or 41 or 42 or 43 or 44
46. 4 and 24 and 38 and 45

**PsycINFO**

1. Obesity/
2. Overweight/
3. obes*.tw.
4. overweight.tw.
5. 1 or 2 or 3 or 4
6. weight loss/ or weight control/

| |
|---|
| 7. diets/<br>8. exp bariatric surgery/<br>9. exp exercise/<br>10. (diet* adj2 therap*).tw.<br>11. bariatric*.tw.<br>12. (low adj3 (energy or calor*) adj4 diet).tw.<br>13. ((pharma* or diet* or obes* or lifestyle or behavio*) adj3 (interven* or treat* or therap*)).tw.<br>14. ((calori* or diet*) adj3 (reduc* or restrict*)).tw.<br>15. (weight adj3 (manag* or los*)).tw.<br>16. exercis*.mp. or physical activit*.tw.<br>17. HAES.mp.<br>18. health at every size.mp.<br>19. (weight adj2 neutral).mp.<br>20. nondiet.mp.<br>21. (non adj2 diet).mp.<br>22. (intuitive adj2 eat*).mp.<br>23. mindful*.tw.<br>24. 6 or 7 or 8 or 9 or 10 or 11 or 12 or 13 or 14 or 15 or 16 or 17 or 18 or 19 or 20 or 21 or 22 or 23<br>25. Body Image/<br>26. (body adj3 (accept* or dissatisf* or image or satisf* or appreciat* or esteem)).tw.<br>27. eating disorders/ or anorexia nervosa/ or binge eating disorder/ or bulimia/ or hyperphagia/ or "purging (eating disorders)"/<br>28. eating behavior/ or binge eating/ or dietary restraint/<br>29. (bulimi* adj3 symptom*).tw.<br>30. (disorder* adj3 eat*).tw.<br>31. (emotion* adj3 eat*).tw.<br>32. (diet* adj3 restr*).tw.<br>33. (binge adj3 eat*).tw.<br>34. extreme weight loss.tw.<br>35. loss of control.tw.<br>36. drive for thinness.tw.<br>37. ((weight or shape or eat*) adj3 concern).tw.<br>38. 25 or 26 or 27 or 28 or 29 or 30 or 31 or 32 or 33 or 34 or 35 or 36 or 37<br>39. randomized controlled trials/ or clinical trials/ or randomized clinical trials/<br>40. (randomi?ed controlled trial* or RCT* or (controlled adj3 trial)).mp.<br>41. randomi?ed.ti.<br>42. randomly.ab.<br>43. trial.mp.<br>44. clinical trial.mp.<br>45. 39 or 40 or 41 or 42 or 43 or 44<br>46. 5 and 24 and 38 and 45 |
| **SCOPUS**<br>( TITLE-ABS-KEY ( "clinical trials" OR "clinical trials as a topic" OR "randomized controlled trial" OR "Randomized Controlled Trials as Topic" OR "controlled clinical trial" OR "Controlled Clinical Trials as Topic" OR "Clinical trial*" OR trial* OR rct OR random* ) ) AND ( ( TITLE-ABS-KEY ( obes* OR overweight* ) ) AND ( ( TITLE-ABS-KEY ( "Weight loss" OR diet* OR bariatric* OR exercis* OR "anti-obesity agent*" OR haes OR "health at every size" OR "Weight neutral" OR "Intuitive eat*" OR mindful* ) ) OR ( TITLE-ABS-KEY ( ( ( pharma* OR diet* OR obes* OR lifestyle OR behavio* ) W/4 ( interven* OR treat* OR therap* ) ) ) ) ) AND ( ( TITLE-ABS-KEY ( ( ( weight OR shape OR eat* ) W/3 concern* ) ) ) OR ( ( TITLE-ABS-KEY ( "Body image*" OR "Eating disorder*" OR anorexia OR "binge eating disorder*" OR bulimi* OR "Emotion* eat*" OR "Diet* restr*" OR "Binge eat*" |

| |
|---|
| OR "extreme weight loss*" OR "loss of Control" OR "Drive for thinness" ) ) OR ( TITLE-ABS-KEY ( ( ( weight OR shape OR eat* ) W/3 concern* ) ) ) ) ) ) |
| **Clinicaltrials.gov**<br>Key search terms via basic search platform:<br>(weight management OR obesity treatment) |
| **WHO ICTRP**<br>Key search terms via basic search platform:<br>(weight management OR obesity treatment) |

**Table S2: Randomised controlled trails eligible for inclusion in the Eating Disorders In weight-related Therapy (EDIT) Collaboration**

This table includes all trials that we have identified up to August 2022 as being eligible for inclusion in EDIT. Trials shaded in blue have agreed to join the Collaboration and share data. Trials shaded in yellow no longer have individual participant data available to share.

| Country/ citation | Trial registration number | Primary contact | Start/ end recruitment period (years) | Sample size | Population | Eating disorder tool/s |
|---|---|---|---|---|---|---|
| **ADOLESCENT TRIALS** | | | | | | |
| Australia, Bonham et al. 2017(1) | ISRCTN13602313 | Maxine Bonham & Aimee L. Dordevic | 2013/2015 | 74 | 13-17y, BMI z-score ≥1.282, >85th percentile | ChEDE, EDE-Q |
| Australia, Brennan et al. 2012(2) | ACTRN12610000111077 | Leah Brennan | | 63 | 11-19y, overweight or obesity | EDI-2 |
| Australia, Lister et al. 2020(3) | ACTRN12617001630303 | Natalie B Lister | 2018/2023 | 186 | 13-17y, adult equivalent BMI > 30 kg/m$^2$ | EDE-Q, BES |
| Australia, Partridge et al. 2020(4) | ACTRN12619000389101 | Stephanie Partridge | 2020/2022 | 150 | 13 -18y, adult equivalent BMI 25.0-29.9 kg/m$^2$ | EDE-Q |
| Australia, Williams et al. | ACTRN12611000139976 | Joanne Williams | 2011-2014 | 570 | 12-17 years; >= 85th centile for age and gender specific BMI data. | EDE-Q |
| Belgium, Braet et al. 2000(5) | | Caroline Braet | 1989/1991 | 136 | 7-17y, seeking treatment for obesity | EDI |
| Belgium, Braet et al. 2004(6) | | Caroline Braet | 1996/1999 | 122 | 7-17y, BMI > 95th percentile | EDE, EDI |
| Belgium, Desmet et al. | ISRCTN47384427 | Maurane Desmet | 2021/ongoing | 70 | 12-16 years; >140% overweight. | ChEDE-Q |
| Belgium, Naets et al. 2018(7) | ISRCTN14722584 | Annelies Van Eyck | 2017/2019 | 200 | 8-18y, overweight or obesity | EDE |
| Brazil, Lofrano-Prado & Lo Prado et al.(8) | | Wagner Prado & Mara Lofrano-Prado | | 62 | 13-18y BMI >95th percentile | EAT, BITE |
| Brazil, Lofrano-Prado et al. 2021(9) | | Wagner Prado & Mara Lofrano-Prado | | 74 | 13-18y, BMI z-score ≥ 2.0 | BES, EAT-26, BITE |
| Brazil, Samara Audi et al. | RBR-7bxzh2r | Andrea Samara Audi | 2018/ongoing | 60 | 13-17 years; BMI z-score greater than or equal to 2 | BES |
| Germany, Blüher et al. 2014(10) | DRKS00005299 | Susann Blüher | 2013/2014 | 65 | 8-18 years; BMI > 97th percentile. | ChEDE-Q |
| Netherlands, Jansen et al. 2011(11) | | Elena Jansen | | 98 | ≥130% overweight | EDE-Q |
| Norway, Skjåkødegård et al. 2016(12) | NCT02687516 | Hanna F Skjakodegard & Yngvild S Danielsen | 2014/2018 | 120 | 6-18y, BMI ≥ 35, or ≥ 30 with obesity related co-morbidity | YEDE-Q |

| | | | | | | |
|---|---|---|---|---|---|---|
| Portugal, Ramalho et al. 2020(13) | NCT04642222 | Sofia Ramalho | | 210 | 13-18y, BMI ≥ 85th percentile | ChEAT |
| UK, Croker et al. 2012(14) | ISRCTN 51382628. | Dasha Nicholls | 2004/2008 | 72 | 8-12y, overweight or obesity | ChEAT |
| USA, Boutelle et al. 2011(15) | | Kerri Boutelle | | 36 | 8-12y, BMI ≥ 85th percentile and child eating in the absence of hunger | ChEDE, BES |
| USA, Cardel, Newsome et al. 2022 | NCT04484831 | Michelle Cardel & Faith Newsome | 2020/2021 | 40 | 14-19y, BMI ≥85th percentile for sex and age | EAT-26 |
| USA, Darling et al. 2021(16) | NCT02426436 | Katherine Darling | | 66 | 13-17y, BMI >85th percentile and an absolute BMI <50 $kg/m^2$ | EDE-Q |
| USA, DeBar et al. 2012(17) | NCT01068236 | Lynn DeBar | 2005/2009 | 208 | 12-17y, BMI ≥ 90th percentile | QEWP-Adolescent |
| USA, Douglas et al. 2020(18) | NCT04027426 | Hollie Raynor | 2019 | 156 | 8-12 years, ≥ 85th percentile BMI, one adult caregiver with a BMI ≥ 25 kg/m2. | KEDS |
| USA, Doyle et al. 2008(19) | | Andrea Goldschmidt & Angela Doyle | 2003/2005 | 80 | 12-18y, overweight, ≥85th percentile | EDE-Q |
| USA, Eichen et al. 2019(20) | NCT01197443 | Dawn Eichen & Kerri Boutelle | | 150 | 8-12y, BMI percentile between 85 and 99.9), one parent with BMI >25 $kg/m^2$ | EDE-Q |
| USA, Epstein et al. 2001(21) | | Denise E Wilfley | | 47 | 8-12y, 20% - 100% overweight compared to 50th percentile | BES, KEDS |
| USA, Estabrooks et al. 2009(22) | NCT00433901 | Paul Estabrooks | 2004/2006 | 220 | 8–12y, BMI ≥ 85th percentile | KEDS |
| USA, Follansbee-Junger et al. 2010(23) | | Katherine Follansbee-Junger | 2006/2008 | 68 | 8-13y, overweight or obesity | ChEAT |
| USA, Goldschmidt et al. 2014(24) | | Denise E Wilfley | | 150 | 7-12y, 20–100% above median BMI, with at least one parent with BMI≥25 $kg/m^2$ | ChEDE, EDE-Q |
| USA, Jelalian et al. 2006(25) | | Elissa Jelalian | 2000/2002 | 76 | 13-16y, 20- 80% overweight | BES |
| USA, Raynor et al. 2021*(26) | NCT02586090 | Melanie Bean | 2016/2018 | 82 | 12-16y, BMI ≥ 85th percentile | EDE-Q |
| USA, Rhee et al. | NCT02976636 | Kay (Kyung) Rhee | 2017/2022 | 160 | 7 - 12 y, BMI ≥ 85th but <100% overweight; at least one parent with (BMI≥25) | |
| USA, Saelens et al. 2002(27) | | Brian Saelens | | 44 | 12-16y, 20% - 100% above the median (50th percentile) | ChEAT |

| | | | | | | |
|---|---|---|---|---|---|---|
| USA; Sato et al. | NCT04038684 | Amy Sato | | 240 | 13-18 years, ≥ 85th%ile for age and sex | YEDE-Q |
| USA, Shomaker et al. 2017(28) | NCT00263536 | Marian Tanofsky-Kraff & Jack Yanovski | 2012/2014 | 29 | 8-13y, BMI ≥ 85th percentile, one parent with overweight/ obesity (BMI ≥ 25 kg/m$^2$) | ChEDE, QEWP-Adolescent |
| USA, Vidmar et al. 2020(29) | NCT03954223 | Alaina Vidmar | | 60 | 14-18y, BMI ≥ 95th percentile | BEDS-7 |
| **ADULT TRIALS** | | | | | | |
| Australia, Cheng et al. 2014(30) | ACTRN12609000307202 | Hoi Lun Cheng | 2006/2009 | 71 | 18–25y, BMI ≥ 27.5 kg/m$^2$ | BES |
| Australia, Raman et al. 2018(31) | ACTRN12613000537752 | Evelyn Smith | 2013/2014 | 80 | 18-55y, BMI > 30 kg/m$^2$, weight < 180 kg | EDE-Q |
| Australia, Rieger et al. 2017(32) | ACTRN12611000509965 | Elizabeth Rieger | 2010/2013 | 201 | 18-65y, BMI ≥ 30 kg/m$^2$ | BES, EAT |
| Australia, Seimon et al. 2019(33) | 12612000651886 | Amanda Salis | 2013/2016 | 101 | 45-65y, postmenopausal women, BMI 30-40 kg/m$^2$, sedentary | EDE, EDE-Q |
| Australia, Smith et al. 2017(34) | ACTRN12616000658415 | Evelyn Smith | | 176 | 18–55y, BMI ≥ 30.0 kg/m$^2$ | EDE-Q |
| Australia, Zwickert et al. 2016(35) | | Elizabeth Rieger | 2013/2015 | 60 | 18-65y, BMI ≥ 30.0 kg/m$^2$ | BES |
| Brazil, Bolognese et al. 2020(36) | RBR-2YZS7 | Braulio Henrique Magnani Branco | | 74 | 40–59y, BMI ≥ 25 kg/m$^2$, female | EAT-26 |
| Brazil, Salvo et al. 2018(37) | NCT02893150 | Vera Salvo & Marcelo Demarzo | | 240 | 18-60y, BMI 25-40 kg/m$^2$, female | BES, EAT |
| Canada, Moss et al. 2017(38) | NCT02649634 | Kristin von Ranson | 2007/2010 | 135 | BMI >= 25 kilograms, 18 Years+ | EDE-Q |
| Canada, Tanco et al. 1998(39) | | Wolfgang Linden | | 62 | ≥19y, BMI ≥ 30 kg/m$^2$, female | EDI |
| Finland, Fogelholm et al. 1999(40) | | Mikael Fogelholm | | 85 | 29-46y, premenopausal, BMI 29-46 kg/m$^2$, female | BITE |
| Finland, Keränen et al. 2009(41) | | Anna-Maria Keranen | 2002/2004 | 82 | 18–65y, BMI > 27 kg/m$^2$ | BES |
| Germany, Hilbert et al. 2016(42) | DRKS00005182 | Anja Hilbert | 2013/2017 | 72 | ≥18y, BMI 25-45kg/m$^2$ | EDE-Q8 |
| Greece, Christaki et al. 2013(43) | UoAMedPR-4716-180211-25 | Christina Darviri | 2010/2011 | 34 | BMI > 28 kg/m$^2$, female | EAT-26 |
| Italy, Dalle Grave et al. 2013(44) | USL22#01/07-CEP31 | Riccardo Dalle Grave & Simona Calugi | | 88 | 18-65y, BMI ≥ 40.0 kg/m$^2$ or 35-39.9 kg/m$^2$ with at least one comorbidity | BES |

| | | | | | | |
|---|---|---|---|---|---|---|
| Italy, Muggia et al. 2014(45) | NCT01686854 | Chiara Muggia | 2007/2011 | 163 | 18-65y, BMI 25-39.9 kg/m$^2$ | BITE 16 |
| New Zealand, Jospe et al. 2017(46) | ACTRN12615000010594 | Rachael W. Taylor | 2014/2015 | 250 | ≥18y, BMI > 27 kg/m$^2$ | EDE-Q |
| Netherlands, Boh et al. 2016(47) | NTR5473 | Bastiaan Boh | 2015/na | 134 | 18-60y, BMI > 25 kg/m$^2$ | EDE-Q |
| Netherlands, Dassen et al. 2018(48) | | Katrijn Houben | | 91 | 18-60y, BMI > 25 kg/m$^2$ | EDE-Q |
| Netherlands, Schyns et al. 2020(49) | | Ghislaine Schyns | | 45 | 18-60y, BMI > 27 kg/m$^2$, female | EDE-Q |
| Netherlands, Werrij et al. 2009(50) | | Marieke Werrij | | 204 | 18-65y, BMI > 27 kg/m$^2$ | EDE-Q |
| Romania; Podina et al.(51) | ISRCTN70907354 | Ioana Podina | | 74 | 18-35 years, BMI 25-29.9 kg/m2 | Eating Disorders Beliefs Questionnaire |
| Spain, Félix et al. | NCT03937167 | Miriam Félix | 2017/2019 | 180 | 18-65 years. BMI >= 30 | EDI |
| UK, Beaulieu et al. 2020(52) | NCT03447600 | Kristine Beaulieu | 2018/2018 | 46 | 18-55y, BMI 25.0-34.9 kg/m$^2$ | BES |
| UK, Cooper et al. 2010(53) | | Zafra Cooper | | 150 | 20-60y, BMI 30.0 - 39.9 kg/m$^2$, female | EDE |
| UK, Scott et al. 2019(54) | ISRCTN88405328 | James Stubbs | 2017/2019 | 1627 | ≥18y, BMI > 25 kg/m$^2$, <150kg | BES |
| UK, Simpson et al. 2015(55) | ISRCTN35774128 | Sharon Simpson | 2011/2012 | 170 | 18–70y, BMI of ≥ 30 kg/m$^2$ | EDE-Q |
| UK, Whitelock et al. 2019(56) | NCT03602001 | Victoria Whitelock & Eric Robinson | 2017/20118 | 107 | 18–65y, BMI ≥25 kg/m$^2$ | BES |
| USA, Afari et al. 2019(57) | NCT01757847 | Niloo Afari | | 88 | 18–75y, BMI ≥ 25 kg m$^2$ | BES |
| USA, Ariel et al. 2016(58) | | Aviva H. Ariel | | 612 | 21-75 years, BMI ≥ 30 & ≤ 45 | BES |
| USA, Bacon et al. 2002(59) | | Lindo Bacon | | 78 | 30-45y, BMI 30-45 kg/m$^2$, female | EDI-2 |
| USA, Barnes et al. 2017(60) | NCT02578199 | Rachel Barnes | | 59 | BMI 25-55 kg/m$^2$ | EDE, EDE-Q |
| USA, Boutelle et al. 2019(61) | | Kerri Boutelle | 2015/2017 | 271 | 18–65y, BMI 25-45 kg/m$^2$ | EDE, EDE-Q, BES |
| USA, Carels et al. 2014(62) | | Robert Carels | | 59 | BMI ≥ 27 kg/m$^2$ | BES |
| USA, Carels et al. 2019(63) | | Robert Carels | | 53 | ≥18y, BMI ≥ 27 kg/m$^2$ | BES |
| USA, Carpenter et al. 2019(64) | | Kelly Carpenter | 2014/2015 | 75 | ≥18y, BMI 25-35 kg/m$^2$ | BES |

| | | | | | | |
|---|---|---|---|---|---|---|
| USA, Dennis et al. 1999(65) | | Karen Dennis | | 39 | Obesity | BES |
| USA, Dennis et al. 2001(66) | | Karen Dennis | | 59 | 50-65y, BMI 27-40 kg/m$^2$ | BES |
| USA, DiMarco et al. 2009(67) | | Ilyse DiMarco | | 39 | 18-55y, BMI 27 – 40 kg/m$^2$ | EDE-Q |
| USA, Eichen et al. | NCT03724396 | Dawn Eichen | 2020/2021 | 67 | 18-65y, BMI >25 and ≤45 | EDE, EDE-Q |
| USA, Glynn et al. 2022(68) | NCT03057873 | Erin L Glynn | 2017/2018 | 206 | 25-50 y, BMO 27-35 inclusive, sans childbearing potential | BES |
| USA, Goodrick et al. 1998(69) | | John Foreyt | | 219 | 25-50y, 14 - 41 kgs overweight | BES |
| USA, Hilderbrandt et al. | NCT04797169 | Tom Hildebrandt | 2021/2024 (predicted) | 600 | 18-60 years, BMI > 27 kg/m2 & interested in the Noom app. | EDE-Q |
| USA, Jeffery et al. 1995(70) | | Robert Jeffrey | | 122 | 120-140% of ideal weight | BES |
| USA, Jeffery et al. 1998(69) | | Robert Jeffrey | | 193 | 25-55y, 14 - 32kg overweight | Gormally Binge Eating Questionnaire |
| USA, Kalarchian et al. 2013(71) | NCT00623792 | Melissa Kalarchian | | 240 | ≥18y, seeking bariatric surgery | EDE |
| USA, LaRose et al. 2014(72) | NCT01096719 | Hollie Raynor | | 178 | ≥21y, BMI 27–45 kg/m$^2$ | EDDS |
| USA, Lillis et al. 2015(73) | NCT01461421 | Jason Lillis | | 160 | 18–70y, BMI 25–50 kg/m$^2$ | EDE-Q |
| USA, Martin et al. 2019(74) | NCT01264406 | Corby K Martin | 2010/2015 | 198 | BMI 25-45 kg/m$^2$, sedentary | MAEDS |
| USA, Mason et al. 2019(75) | NCT00470119 | Anne McTiernan | | 439 | 50–75y, BMI ≥25.0 kg/m$^2$ (if Asian-American BMI ≥23.0 kg/m$^2$) Female only. | BES |
| USA, Mensinger et al. 2016(76) | NCT00769717 NCT00769717 | Janell Mensinger | 2008/2011 | 80 | 30-45y, BMI 30 - 45 kg/m$^2$ | EDE-Q |
| USA, Napolitano et al. 2017(77) | NCT02342912 | Melissa A Napolitano | 2015/2017 | 450 | 18–35y, BMI of 25–45 kg/m$^2$ | EDDS |
| USA, Pacanowski et al. 2014(78) | | Carly Pacanowski & Nancy Sherwood | 2007/2008 | 419 | 19-70y, achieved ≥ 10% weight loss | EDDS binge eating items |
| USA, Radin et al. 2020(79) | https://aspredicted.org/nk4av.pdf; #8472 | Rachel Radin | | 194 | ≥18y, BMI 30-45.9 kg/m$^2$ | BES |
| USA, Ramirez et al. 2001(80) | | Elena Ramirez | | 65 | ≥18y, BMI > 27.3 kg/m$^2$ for women, > 27.8 kg/m$^2$ for men | EDE-Q |

| | | | | | | |
|---|---|---|---|---|---|---|
| USA, Raynor et al. 2006(81) | | Hollie Raynor | | 30 | BMI 25 – 40 kg/m$^2$ | BES |
| USA, Smith et al. 2018(82) | NCT02753972 | Brian M Shelley | 2006/2006 | 36 | 50–70y, postmenopausal, BMI > 30 kg/m$^2$ | BES |
| USA, Steinberg et al. 2014(83) | | Dori Steinberg | 2011/2011 | 91 | 18–60y, BMI 25–40 kg/m$^2$ | QEWP-R, Mizes Anorectic Cognitions Questionnaire |
| USA, Stice et al. | NCT03375853 | Eric Stice | 2017/2023 (predicted) | 180 | 18-38 years, BMI 25-35 | EDE |
| USA, Vander et al. 2006(84) | | Nikhil V Dhurandhar | | 80 | 18-65y, BMI ≥ 30 kg/m$^2$ | NESQ |
| USA, Varady et al. | NCT04692532 | Krista Varady | 2021/2023 | 90 | 18-70y, obesity | MAEDS |
| USA, Wadden et al. 1994(85) | | Thomas Wadden | | 49 | ≥25kg overweight | BES |
| USA, Wadden et al. 2004(86) | | Thomas Wadden | | 123 | BMI 30–43 kg/m$^2$, female | EDE |
| USA, Williamson et al. 2008(87) | | Corby K Martin | | 48 | 25-50y, BMI 25-30 +kg/m$^2$ | MAEDS |

Abbreviations: BEDS-7, Binge Eating Disorder Screener-7; BES, Binge Eating Scale; BITE, Bulimic Investigatory Test of Edinburgh; EAT, Eating Attitudes Test; EDDS, Eating Disorder Diagnostic Scale; EDE, Eating Disorder Examination; EDE-Q, Eating Disorder Examination Questionnaire; EDI, Eating Disorder Inventory; MAEDS, Multidimensional Assessment of Eating-Disorder Symptom; NESQ, Night Eating Syndrome Questionnaire; QEWP, Questionnaire of Eating and Weight Patterns.

* Trial includes adolescents and adult data

**Table S3: Funding sources for trials which have agreed to join the EDIT Collaboration**

| Trial Author, year of publication | Funding |
|---|---|
| ADOLESCENT TRIALS | |
| Bonham et al. 2017 | Jenny Craig Weight Loss Centers Pty Ltd (to M.B. and H.T.). The funders had no role in the design and conduct of the study; collection, management, analysis, and interpretation of the data; and preparation, review, or approval of the manuscript; nor the decision to submit the manuscript for publication. However, employees of Jenny Craig (consultants) were involved with the initial recruitment of participants and collection of anthropometric data. |
| Boutelle et al. 2011 | University of Minnesota Faculty Development Grant to Kerri N. Boutelle and Lisa Harnack. |
| Braet et al. 2004 | N/A |
| Cardel, Newsome et al. 2022 | National Institute of Health National Heart, Lung, and Blood Institute K01HL141535. |
| Croker et al. 2012 | Cancer Research UK, Great Ormond Street Hospital and Weight Concern. |
| Douglas et al. 2020 | National Institutes of Health under award 1R01DK121360–01. |
| Eichen et al. 2019 | National Institute of Diabetes and Digestive and Kidney Diseases, Grant/Award R01DK075861. |
| Epstein et al. 2001 | N/A |
| Goldschmidt et al. 2014 | NIH grants R01 HD036904, K24 MH070446, and T32 HL007456, and NCRR grants KL2 RR025000, KL2 RR024994, and UL1 RR024992. This research was supported in part by a Student Research Award by the American Psychological Association's Division 38 (Health Psychology) to Angela Celio Doyle, Ph.D., grant 1K24MH070446-02 from the National Institute of Mental Health, National Institutes of Health, awarded to Denise E. Wilfley, Ph.D., and an RGA/Washington University Longer Life Foundation Research Award to Dr. Wilfley. |
| Lister et al. 2020 | Australian National Health and Medical Research Council (NHMRC) project grant funding 2017–2020 (#1128317). |
| Lofrano-Prado & Prado et al. 2017 | National Council for Scientific and Technological Development - CNPQ and Foundation for Science and Technology of the State of Pernambuco – FACEPE. |
| Lofrano-Prado et al. 2021 | Coordination for the Improvement of Higher Education Personnel (CAPES). |
| Partridge et al. 2020 | NSW Health Early-Mid Career Researcher Grant under the NSW Cardiovascular Research Capacity Program. |
| Ramalho et al. 2020 | Research was partially conducted at the Psychology Research Centre (PSI/01662), University of Minho, and supported by the Portuguese Foundation for Science and Technology and the Portuguese Ministry of Science, Technology and Higher Education through national funds, and co-financed by FEDER through COMPETE2020 under the PT2020 Partnership Agreement (POCI-01-0145-FEDER-007653), by the following grants to Eva Conceição (IF/01219/2014 and POCI-01-0145-FEDER-028209), and doctoral scholarship to Sofia Ramalho (SFRH/BD/104182/2014). The work from Pedro Saint-Maurice was partially funded by an individual fellowship grant awarded by the Fundação para a Ciência e a Tecnologia (FCT; Portugal) (SFRH/BI/114330/2016) under the POPH/FSE program. |
| Raynor et al. 2021 | National Institute of Child Health and Human Development (R21HD084930) awarded to MKB. Additional support was obtained from the Children’s Hospital Foundation (unnumbered, to MKB and EPW) and the National Center for Advancing Translational Science (CTSA award UL1TR002649 to FGM). These funding agencies had no influence over the design or conduct of this work. |

| | |
|---|---|
| Shomaker et al. 2017 | National Institutes of Health Intramural Research Program Grant 1ZIAHD000641 from NICHD (JAY). Pilot Intramural Research Award 72ON-01 from USUHS (MTK). |
| Skjåkødegård et al. 2016 | Health Authorities of Western Norway and the University of Bergen. Accelerometers used in the trial were funded by a grant from the non-commercial organization Sov.no (Norwegian Competency Service for Sleep Disturbances). No commercial companies were funding any salaries or equipment for this trial. |
| Vidmar et al. 2020 | This work was supported in part by grants (1) UL1TR001855 from the National Center for Advancing Translational Science (NCATS) of the U.S. National Institutes of Health, (2) NIH/NCRR SC-CTSI Grant Number UL1 TR000130, (3) National Institute on Minority Health and Health Disparities (NIMHD) Obesity Health Disparities Research Center (U54MD000502; Salvy/Dutton), the Eunice Kennedy Shriver National Institute of Child Health and Human Development (NICHD; R01HD092483; de la Haye/Salvy), and (4) the National Cancer Institute (NCI; 1R01CA258222, Figueiredo/Salvy/Peterson). The content is solely the responsibility of the authors and does not necessarily represent the official views of the National Institutes of Health. Dexcom supported this study by providing the continuous glucose monitor equipment required. |
| **ADULT TRIALS** | |
| Barnes et al. 2017 | NIH career development awards, K23-DK092279 for RDB and K24-DK070052 for CMG. |
| Beaulieu et al. 2020 | Funded by a Research Fellowship awarded to Kristine Beaulieu by the European Society for Clinical Nutrition and Metabolism (ESPEN) and in-kind support from LighterLife UK Ltd. The funding sources had no involvement in the conduct of the research and preparation of the article. |
| Bolognese et al. 2020 | N/A |
| Boutelle et al. 2019 | National Institutes of Health [R01DK103554, K23DK114480, UL1TR001442]. The content is solely the responsibility of the authors and does not necessarily represent the official views of the National Institutes of Health. |
| Carpenter et al. 2019 | NCCIH/NIH grant 1R21AT007845-01A1. K Carpenter PI. |
| Cheng et al. 2014 | Meat and Livestock Australia (Project No. D.MHN.0009) awarded to Dr Helen O'Connor. Meat and Livestock Australia had no role in the study design, collection, analysis, interpretation of data or writing of the paper including the decision to submit for publication |
| Dalle Grave et al. 2013 | N/A |
| Dassen et al. 2018 | Maastricht University Interfaculty Program 'Eatwell' |
| Hilbert et al. 2016 | Grant 01EO1001 from the German Federal Ministry of Education and Research. |
| Jospe et al. 2017 | SWIFT was funded the University of Otago (no grant number) |
| LaRose et al. 2014 | R01DK074721 from the National Institute of Diabetes and Digestive and Kidney Diseases to HAR and K23DK083440 from the National Institute of Diabetes and Digestive and Kidney Diseases to JGL. |
| Martin et al. 2019 | Supported by the NIH via the National Heart, Lung, and Blood Institute, with the Multiple Principal Investigators being CKM and TSC (R01 HL102166); NORC Center grant P30 DK072476, titled "Nutritional Programming: Environmental and Molecular Interactions," sponsored by the National Institute of Diabetes and Digestive and Kidney Diseases; the National Institute of General Medical Sciences, which funds the Louisiana Clinical and Translational Science Center (U54 GM104940); and NIH grant F32 HL123242. Grant R01 HL102166 was the main funding source for the project. |
| Mason et al. 2019 | Funded in part through the NIH/NCI Cancer Center Support Grant P30 CA015704, and supported by grants from the National Cancer Institute at the National Institutes of Health: R01 CA105204-01A1, NIHR03 CA162482–01 and grants from the Breast Cancer Research Foundation (BCRF-16-106 and BCRF-17-105). The funding bodies had no role in the |

| | |
|---|---|
| | design of the study nor in the collection, analysis and interpretation of data, or manuscript preparation. |
| Mensinger et al. 2016 | Grant awarded to the first author from the Edna G. Kynett Memorial Foundation. The Reading Hospital and Medical Center sponsored the project by holding the research at their site and paying the salary and benefits of the PI and Research Coordinators that went above and beyond the 85k in funding from the Kynett Foundation. |
| Pacanowski et al. 2014 | Grant from the NIH/National Cancer Institute (R01 CA128211) and CP was supported by a University of Minnesota training grant (NIH T32 DK083250). |
| Raman et al. 2018 | Diabetes Australia Research Trust Grant 2014. |
| Reiger et al. 2017 | National Health and Medical Research Council of Australia project grant (#632621). |
| Salvo et al. 2018 | Biochemistry examinations will be conducted by the São Paulo Municipal Health Department (in BHUs) and the remainder will be performed using own resources. The Brazilian Centre of Mindfulness and Health Promotion (Mente Aberta Centre) for subsidising the materials needed for this research (photocopy, electrodes for bioimpedance). |
| Seimon et al. 2019 | Project Grant (1026005) from the National Health and Medical Research Council of Australia, awarded to A. Sainsbury, N. Byrne and I. Caterson. The Rebecca L. Cooper Medical Research Foundation and the University of Sydney/National Health and Medical Research Council of Australia provided grants that contributed to the purchase of equipment used for this trial. Prima Health Solutions (Brookvale, New South Wales, Australia) provided inkind support for this trial in the form of below-cost KicStart meal replacement products (shakes) and a gift of associated adherence tools (shakers). This relationship with Prima Health Solutions was established after the dietary protocol for the TEMPO Diet Trial had been established. |
| Simpson et al. 2015 | National Institute for Health Research Health Technology Assessment programme. |
| Smith et al. 2017 | Ramaciotti Foundation Australia (HIG2015/075) and Western Sydney University (Project code: 20311.64321). |
| Smith et al. 2018 | Financial support from the La Tierra Sagrada Society of the University of New Mexico (UNM) and in-kind support from the UNM General Clinical Research Center. |
| Varady et al. | Trial is ongoing |
| Whitelock et al. 2019 | Economic and Social Research Council [project reference: ES/N00034X/1]. The funding body had no role in the design of the trial, data collection, analysis and interpretation, or in writing the manuscript. |
| Williamson et al. 2008 | National Institute on Aging, National Institutes of Health (U01AG022132, U01AG020478, U01AG020487, and U01AG020480). |
| Zwickert et al. 2016 | Supported by the NHMRC via a Senior Research Fellowship to Amanda Sainsbury and by a Program Grant of the NHMRC APP1037786. |
